# Knowledge, attitude and practice of puerperal and expectant mothers on neonatal jaundice in selected health facilities in Tamale metropolis, Ghana

**DOI:** 10.1101/2025.08.13.25333555

**Authors:** Francis Kpornu Cudjoe, Abdul-Samed Mohammed, Daudi Yeboah, Jonathan Mawutor Gmanyami, Vida Nyagre Yakong, Michael Yaw Amoakoh

## Abstract

**Background:** Neonatal jaundice (NNJ) remains a prevalent and potentially dangerous condition during the neonatal period. It is a leading cause of neonatal hospitalization and, if not recognized early, can result in irreversible neurological complications such as kernicterus. Globally, the burden of NNJ is disproportionately high in low- and middle-income countries, especially in the Ghanaian setting. Despite this, community-level awareness and timely care-seeking remain suboptimal due to myths, misconceptions, and inadequate knowledge among mothers.

**Objective:** To assess the knowledge, attitude, and practices of puerperal and expectant mothers regarding neonatal jaundice in Tamale Metropolis.

**Methods:** A descriptive cross-sectional study was conducted among 229 puerperal and expectant mothers recruited from three public health facilities using multistage random sampling. Structured questionnaires were used to assess knowledge, attitude, and practices. Data were analyzed using SPSS version 26. Descriptive statistics and multivariate regression were used to examine associations between sociodemographic variables and outcomes. Statistical significance was determined at p<0.05.

**Results:** Among 229 participants, 50.7% were aged 25–35 years. Overall, 55.0% demonstrated good knowledge of NNJ, while 45.0% had poor knowledge. Regarding attitudes, 65.1% showed a positive attitude towards NNJ. Most respondents (78.2%) believed that NNJ requires hospital treatment, and 56.8% endorsed exposing newborns to early morning sunlight. In terms of beliefs, 81.7% rejected the notion that jaundice enhances a baby’s appearance, and 79.9% disagreed that yellow discoloration indicates healthy growth. About 72.0% exhibited good beliefs and practices. Knowledge of mothers, attitude and perception, and employment status were found to be significant determinants of beliefs and practice toward NNJ.

**Conclusion:** While most mothers demonstrated favorable attitudes and appropriate practices towards neonatal jaundice, gaps in knowledge persist. Strengthening health education during antenatal and postnatal care is essential to promote timely recognition and management of NNJ.

## Introduction

Neonatal jaundice (NNJ), characterized by the yellow discoloration of the skin and sclera due to elevated serum bilirubin, remains one of the most frequent clinical conditions requiring attention during the neonatal period [1, 2]. Globally, it affects approximately 60% of term and 80% of preterm neonates within the first week of life, underscoring its pervasiveness and significance [3, 4]. Although many cases resolve without complications, severe or untreated NNJ may lead to irreversible outcomes such as kernicterus and cerebral palsy, particularly in low- and middle-income countries [5, 6].

Despite improvements in neonatal care, sub-Saharan Africa continues to bear a disproportionate burden, accounting for over 75% of global neonatal jaundice-related deaths and disabilities [7, 8]. In Ghana, the trend is similarly troubling. In Ghana, reported cases of neonatal jaundice have steadily risen from 3,031 in 2015 to 9,273 in 2019 [9]. A facility-based study conducted at the Tamale Teaching Hospital in northern Ghana reported that a significant number of neonates with jaundice presented late with elevated bilirubin levels, requiring intensive treatment including phototherapy and, in some cases, exchange transfusion [10].

Poor caregiver knowledge has been linked to delays in seeking appropriate medical treatment, often resulting in severe forms of NNJ by the time care is sought [11]. A study in Ghana reported that 56.2% of mothers believed that neonatal jaundice could be treated at home without hospital care [12]. In addition, cultural beliefs associating jaundice with spiritual causes contribute significantly to underutilization of health services [9, 13]. These misconceptions, combined with inadequate counselling during antenatal care, create systemic gaps in the early detection and management of NNJ [14].

The persistence of severe neonatal jaundice in Ghana is not solely due to biomedical factors but also deeply rooted in socio-cultural beliefs, health system limitations, and caregiver behaviors. Misinformation and traditional practices such as exposing babies to sunlight, using herbal remedies, or delaying care-seeking remain common, even among educated mothers [9]. Furthermore, poor knowledge of danger signs, inappropriate treatment choices, and misattribution of causes to spiritual or maternal failings continue to undermine early intervention efforts [11, 15]. In Tamale and the broader Northern Region of Ghana, NNJ remains among the top five causes of neonatal admissions, with significant proportions of affected newborns presenting late and with severe complications [10, 16]. These observations highlight a pressing need to assess maternal knowledge, attitudes, and perceptions of NNJ within this context, particularly given the role of mothers as primary decision-makers during the early neonatal period. Mothers’ understanding and interpretation of NNJ symptoms directly influence how quickly and appropriately care is sought. In settings like Tamale, where traditional beliefs and limited health literacy may delay clinical intervention, targeted education is essential. Gaining deeper insight into maternal perspectives is therefore critical to developing context-specific, culturally sensitive interventions that promote early recognition, reduce harmful practices, and improve neonatal outcomes in Ghana.

## Material and methods

### Study Design, Area and Population

A descriptive cross-sectional design was adopted for this study. This design was selected because it enables the capture of data from the target population at a single point in time, making it suitable for assessing knowledge, attitudes, and perceptions without the need for extended follow-up. This study was conducted in the Tamale Metropolitan Area, the administrative capital of Ghana’s Northern Region. Located approximately 600 kilometers north of Accra, Tamale is not only the political and commercial hub of the region but also serves as a central point for economic and educational activities in Northern Ghana. The majority of its population identifies as Dagombas and practices Islam. The city’s economy largely depends on trading and farming, and the general educational attainment of residents ranges from basic to junior high school levels. As of 2023, the estimated population of the metropolis stood at 730,000, reflecting a 4.14% increase from the previous year.

The research was carried out in three public health facilities within the metropolis: Tamale Teaching Hospital (TTH), Tamale West Hospital (TWH), and Tamale Central Health Center (TCHC). TTH, located in the Dohinayili community, is the largest and third-oldest teaching hospital in Ghana, serving as a tertiary referral center for the northern regions with over 1,200 nurses. TWH, located in Zogbeli, is another NHIS-accredited primary hospital serving the general population and employing 401 nurses. The fourth facility, TCHC, is positioned along Hospital Road, near the Ola Cathedral and the National Investment Bank, providing general health services within the metropolis.

The study targeted two key groups: expectant mothers attending antenatal care (ANC) and puerperal mothers receiving postnatal care within the selected health facilities. Expectant mothers were defined as those currently pregnant and preparing to deliver, while puerperal mothers referred to those who had delivered a live infant less than six weeks prior to data collection.

### Sample Size Determination and Technique

The sample size was determined using the Taro Yamane formula, which is appropriate for calculating sample sizes when the total population is known. The formula used was: n = N/ 1+ N(e)^2^; where n= required sample size from the population under study; N= the whole population under study; e= margin of error/sampling error.

In this study, the population size NNN was estimated to be 633, which represents the average number of antenatal care (ANC) registrants from October 2023 to December 2023 across the three selected health facilities: Tamale Teaching Hospital (TTH), Tamale West Hospital (TWH), and Tamale Central Health Centre (TCHC) (Table 1). This period was chosen because it reflects a consistent and recent client load relevant to the study’s timeframe.

**Table 1:** Estimated Sample Size for Each Facility.

| **Selected Facilities** | **Last Quarter ANC Registrant from 2023 (October to December 2023)** | **Sampling Size** |
| --- | --- | --- |
| Tamale Teaching Hospital | 683 | 82 |
| Tamale West Hospital | 383 | 46 |
| Tamale Central Health Center | 833 | 101 |
| **Total** | **1899** | **229** |

To determine an appropriate sample size, a margin of error (e) of 5.3% (0.053) was adopted. This level of precision is within the acceptable range for social and health science research, and it strikes a balance between data accuracy and feasibility given the available resources and timeframe. Substituting the values into the formula;

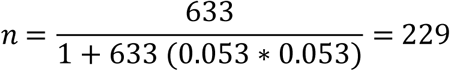

Thus, a sample size of 229 participants was determined to be appropriate for the study. This ensures that the findings are representative of the population while maintaining an acceptable level of precision for drawing inferences.

A multistage sampling technique was used to systematically select participants for this study across three major health facilities in the Tamale Metropolis. This approach was chosen to ensure a balanced, representative sample while reducing the risk of selection bias and enhancing the reliability of findings. In the first stage, purposive sampling was employed to select three public health facilities, namely the Tamale Teaching Hospital (TTH), Tamale West Hospital (TWH), and Tamale Central Health Centre (TCHC). These facilities were selected because they serve as major maternal and neonatal care centres in the metropolis, receiving a high volume of antenatal and postnatal clients, and offering a broad cross-section of the target population for the study.

In the second stage, proportional allocation was applied to distribute the total sample size of 229 among the three facilities (Table 1). This was based on each facility’s total ANC registrants for the last quarter of 2023 (October to December 2023) (DHIMS II, 2024). In the third stage, simple random sampling was used to select eligible mothers from each facility. Upon arrival at the antenatal or postnatal clinic, health records (ANC or PNC books) were reviewed to verify whether the mother met the inclusion criteria. Those who qualified were then asked to participate in a lottery draw. In this process, each eligible mother picked from a bag containing folded pieces of paper marked “Yes” or “No” in equal proportions. Mothers who selected “Yes” were immediately recruited into the study, while those who picked “No” were excluded without prejudice. This method was carried out consecutively on clinic days until the required number of participants from each facility was reached.

This multistage process ensured fairness, reduced sampling bias, and upheld the integrity of the study by giving every eligible mother an equal chance of participating, while also maintaining proportionality across selected facilities.

The proportionate sampling formula is given as;

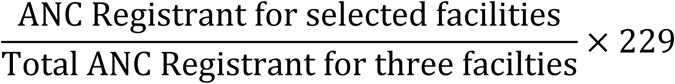

### Study Variables

The main outcome variable for this study was mothers’ traditional beliefs and practices toward neonatal jaundice (NNJ). This variable was assessed through responses to nine (9) items that examined what mothers do when their newborns become jaundiced. Traditional beliefs and practices were captured through items examining spiritual, cultural, and home-based actions taken by mothers to prevent or manage NNJ. These included beliefs in curses, herbal use, avoidance of colostrum, or harmful practices like placing babies in dark rooms. Each of these responses was coded as ‘1’ for correct or non-harmful belief and ‘0’ for harmful or incorrect traditional belief and practice.

The key independent variables included maternal knowledge, attitudes, traditional beliefs, and various socio-demographic characteristics. Maternal knowledge on NNJ was assessed based on correct understanding of the signs, causes, danger signs, complications, and appropriate treatments of NNJ. Responses to each knowledge-related item were coded as ‘1’ for correct and ‘0’ for incorrect or “not sure” responses. Similarly, attitudinal and perceptual variables were assessed through statements gauging maternal disposition toward jaundice prevention, recognition, and treatment. Each attitude-related response was coded as ‘1’ for favorable attitudes (e.g., agreement with hospital-based treatment, ANC education) and ‘0’ for unfavorable or incorrect beliefs (e.g., attributing jaundice to evil spirits, using non-clinical remedies).

The socio-demographic variables included maternal age, marital status, sex, ethnicity, religion, employment status, educational level, parity, ANC visit frequency, gestational trimester, and history of NNJ exposure. These variables were categorized and coded as follows: age was grouped into four categories—18–24, 25–35, 36–45, and 46 years and above; marital status as single, married, divorced, or widowed; sex as male or female; ethnicity into major regional groups with provision for others; and religion into Christian, Muslim, Traditionalist, or others. Employment status was coded as self-employed/private, unemployed/student, public servant, or other. Educational level was categorized as none, primary/basic, secondary, or tertiary. Parity was coded based on number of children: none, one, two, or more than two. Frequency of antenatal care (ANC) attendance was captured as less than four visits or four or more visits, while gestational trimester was coded into first, second, third, or not specific. Respondents were also asked if they had ever received education on NNJ or had a child diagnosed with NNJ, with binary responses coded as ‘1’ for “Yes” and ‘0’ for “No.

### Data Collection Procedure and Instrument

Data were collected between 5^th^ January 2024 to 7^th^ March 2024 at the Antenatal and Postnatal Units of Tamale Teaching Hospital (TTH), Tamale West Hospital (TWH), and Tamale Central Health Centre (TCHC). Eligible participants were approached individually at times convenient to them, and the purpose of the study was clearly explained. After responding to their concerns and obtaining informed consent, data were collected using the KoboCollect mobile application installed on Android tablets.

Each interview was conducted privately within designated spaces to ensure participant confidentiality. Respondents who could read and understand English completed the questionnaire themselves. Those who were unable to read or write in English were assisted by trained research assistants who conducted oral translations. Two members of the research team were fluent in Dagbanli, while others provided translation in Hausa, Twi, and other locally spoken languages to support clear understanding of the questions. Clarifications were given when needed to avoid misinterpretation.

All completed forms were uploaded daily to a secure cloud server, with backups stored under strict password-protected systems. Before leaving the data collection site each day, all entries were reviewed for accuracy and completeness.

The data collection tool used for the study was a self-developed, structured questionnaire designed based on a careful review of existing literature and guided by the study’s objectives. The questionnaire contained a total of 51 questions, including both closed and open-ended items. It was organized into four main sections. Section A gathered information on socio-demographic characteristics, including age, marital status, educational background, ethnicity, number of children, and gestational age. Section B focused on maternal knowledge of neonatal jaundice and included 28 questions on the signs, causes, consequences, and treatment options of the condition. Section C explored maternal attitudes and perceptions with 12 questions, while Section D assessed traditional beliefs and practices with 9 questions that examined culturally rooted responses to neonatal jaundice.

To measure the variables; knowledge, attitude, and practice, a composite scoring system was used based on the responses from Sections B, C, and D of the questionnaire. For each item, a correct response was assigned a score of ‘1’, while an incorrect response was given a score of ‘0’. The cumulative score from each section determined whether a respondent’s level of knowledge, attitude, or practice was considered good or poor. A cumulative score of 80% or higher was categorized as “good,” while a score below 80% was categorized as “poor.” These scores were subsequently coded numerically as ‘1’ for good and ‘0’ for poor to facilitate statistical analysis. This scoring system provided a simple yet effective way of classifying respondents’ understanding and behavior related to neonatal jaundice within the study population.

### Data Analysis Plan

Field data collected were first entered and cleaned using Microsoft Excel to ensure consistency and accuracy before being exported to SPSS version 26.0 for analysis. The data cleaning process involved reviewing the entries for completeness, detecting outliers, identifying inconsistencies, and resolving any discrepancies. These quality checks were essential to prepare the data for robust statistical analysis and reduce errors that could affect the validity of the findings. Descriptive statistics were employed to summarize the socio-demographic characteristics and categorical variables of the study participants. Frequencies and percentages were used to present distributions of key variables such as age, marital status, education level, parity, religion, and ethnicity. These were displayed using simple distribution tables and supported by visual representations such as bar charts and graphs to aid interpretation and clarity.

Multivariate regression analysis was used to determine the associations between the outcome variable which is beliefs and practices and the sociodemographic predictors. All explanatory variables were first examined for multicollinearity, and none met the exclusion threshold of a Variance Inflation Factor (VIF) of 10 or higher, indicating that multicollinearity was not a concern in the regression model. Statistical significance for all associations was determined at a confidence level of 80%, and a p-value less than 0.05 was considered statistically significant.

### Ethical Consideration

Ethical clearance for the study was obtained from the University for Development Studies Institutional Review Board (UDSIRB) with reference number UDS/RB/193/23. In addition to this approval, official permission letters were secured from the administration of the selected health facilities: Tamale Teaching Hospital, Tamale West Hospital, and Tamale Central Health Centre, before the commencement of data collection. These ethical and administrative approvals, along with the detailed research proposal, formed the basis for conducting the study following established ethical standards and institutional requirements.

Verbal consent was obtained from each participant before data collection. Verbal consent was documented in the data collection logbook and witnessed by members of the research team. Before administering the questionnaires, the purpose and relevance of the study were clearly explained to participants in a language they understood to ensure informed decision-making. Participation in the study was entirely voluntary, and respondents were assured that they could withdraw at any stage of the process without facing any consequences or losing access to services. No form of coercion or inducement was employed to influence participation.

Participants were further assured of confidentiality and anonymity throughout the research process. To uphold data privacy, no personal identifiers such as names or contact details were linked to the responses, and the data collected was used strictly for academic purposes. The researchers took every precaution to ensure that participants were not subjected to any form of harm, discomfort, or emotional distress. Data were securely stored and accessible only to the core research team to prevent unauthorized access or disclosure. These measures ensured that the study met ethical standards for the protection of human subjects.

## Results

A total of 229 respondents participated in the study, with the majority aged between 25 and 35 years (50.7%). Most of the respondents were married (55.5%), and 33.2% identified as Dagombas. About half (50.2%) had attained tertiary education, while 31.9% had completed secondary school. In terms of employment, 40.2% were either unemployed or students, and 34.0% were self-employed. The majority (60.7%) had attended four or more antenatal care visits during their pregnancy. More than half (51.1%) reported receiving education on neonatal jaundice, and 20.1% had a child previously diagnosed with the condition (Table 2).

**Table 2:**
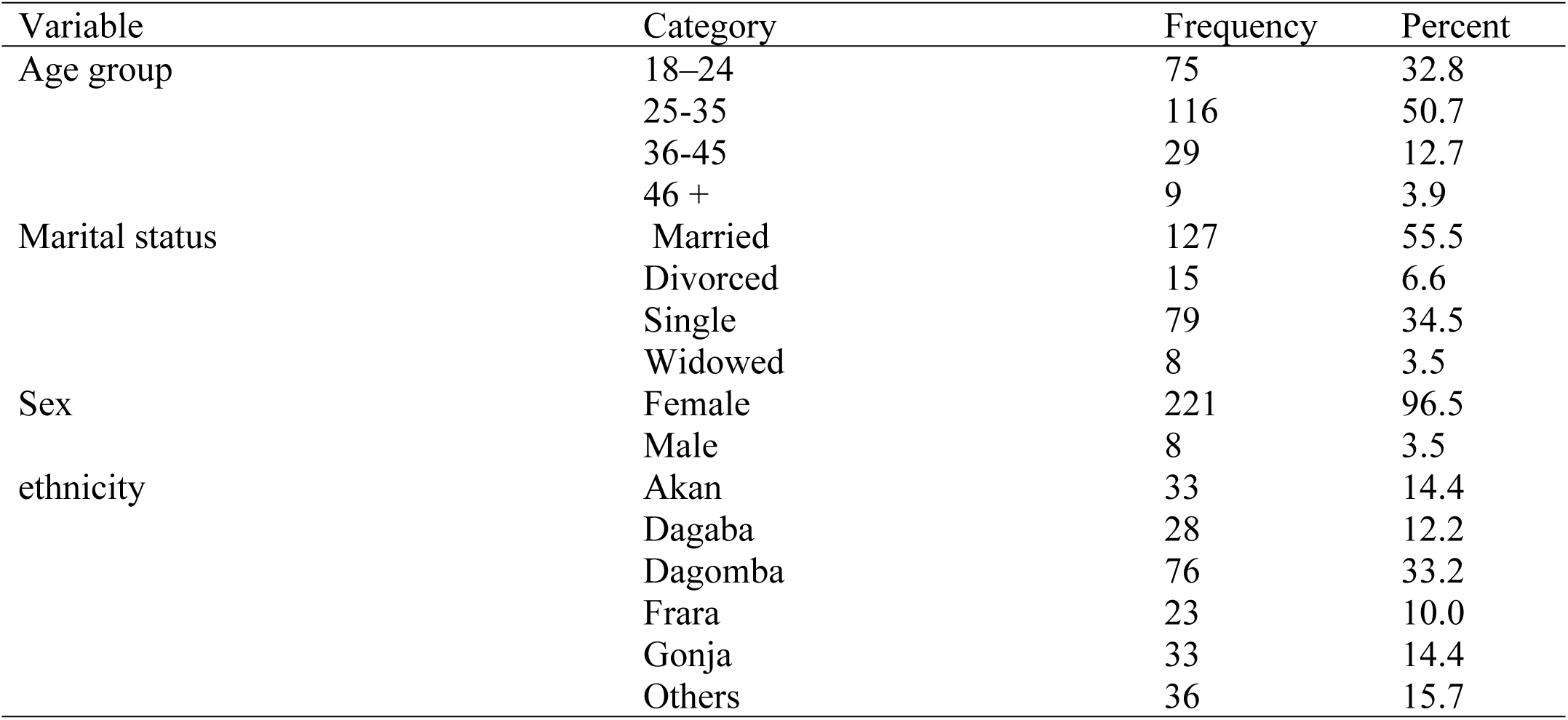
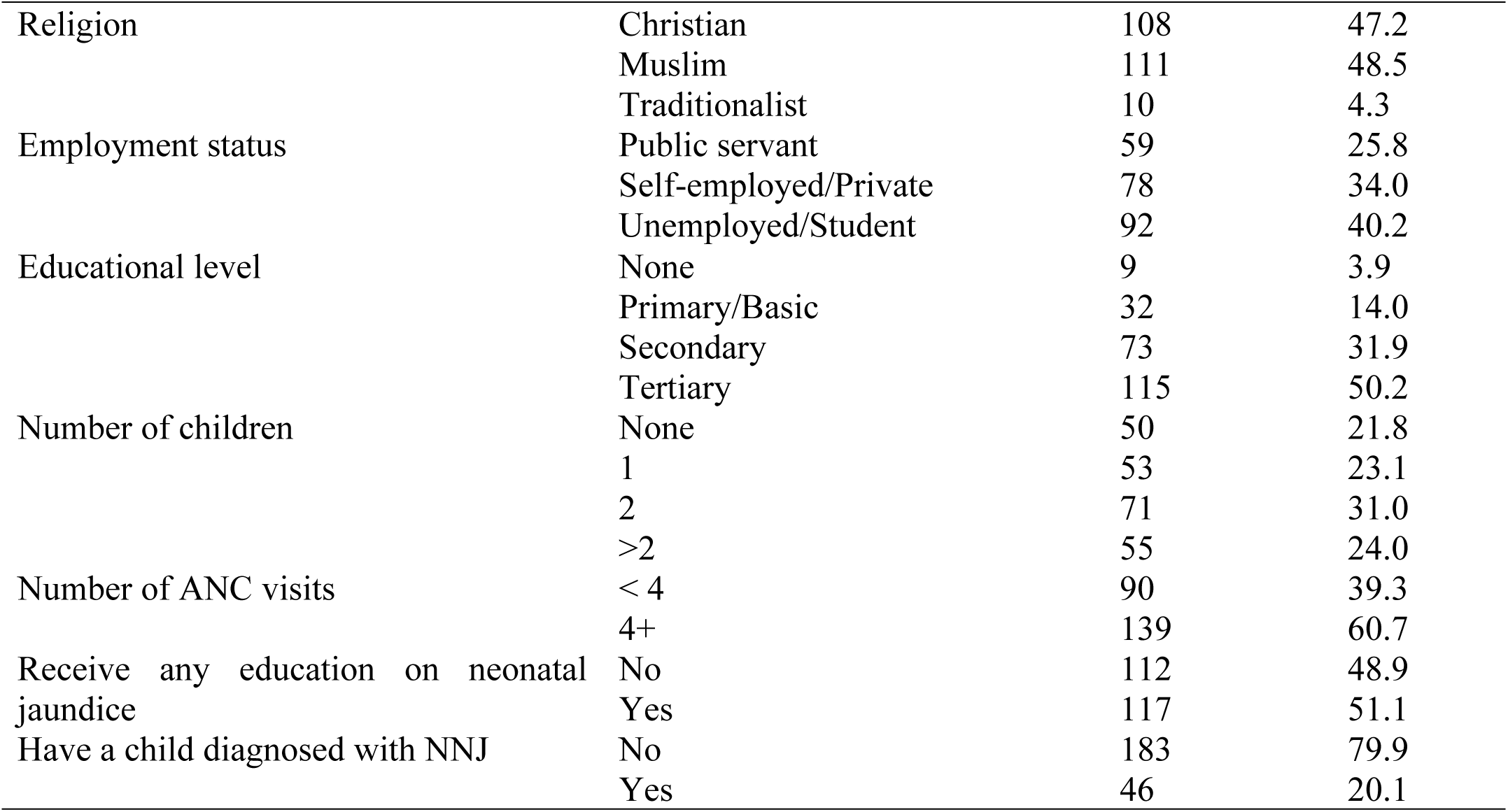
Sociodemographic Characteristics of Mothers.

Out of the 229 respondents, 126 (55.0%) had good knowledge of neonatal jaundice, with the majority receiving their information from health workers (35.4%), followed by mass media (23.1%) (Table 3). The majority of the respondents identified yellowing of the eyes (86.5%) and skin (77.7%) as key signs of neonatal jaundice. Regarding causes, over half (53.7%) mentioned infections, while 42.8% recognized prematurity as a contributing factor. Most respondent noted poor feeding (68.6%), a high-pitched cry (54.6%), and abnormal movement (48.5%) as warning signs. In terms of complications, the possibility of death was the most acknowledged (74.2%) which was followed by brain damage (69.4%). For treatment options, phototherapy was widely recognized by respondents (69.4%) as the main approach.

**Table 3:**
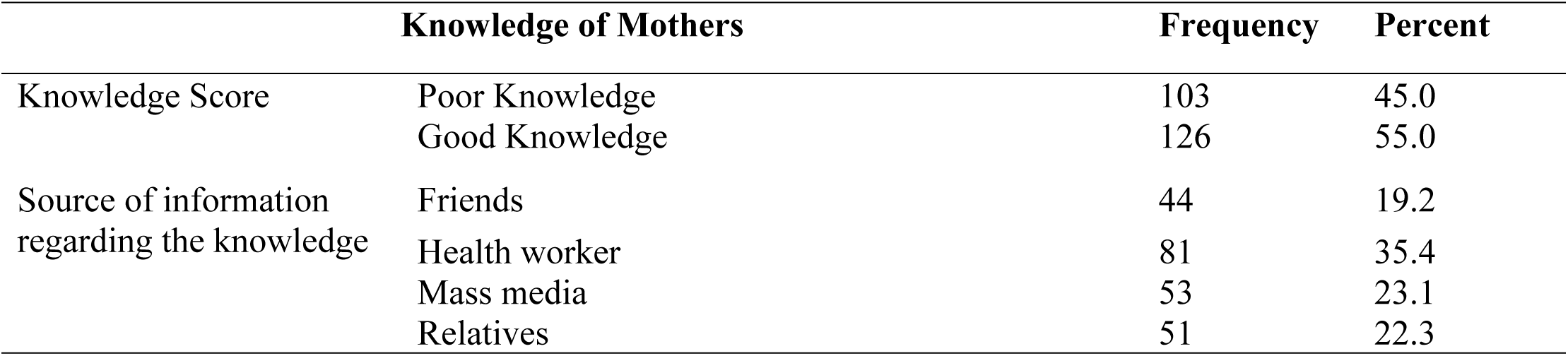
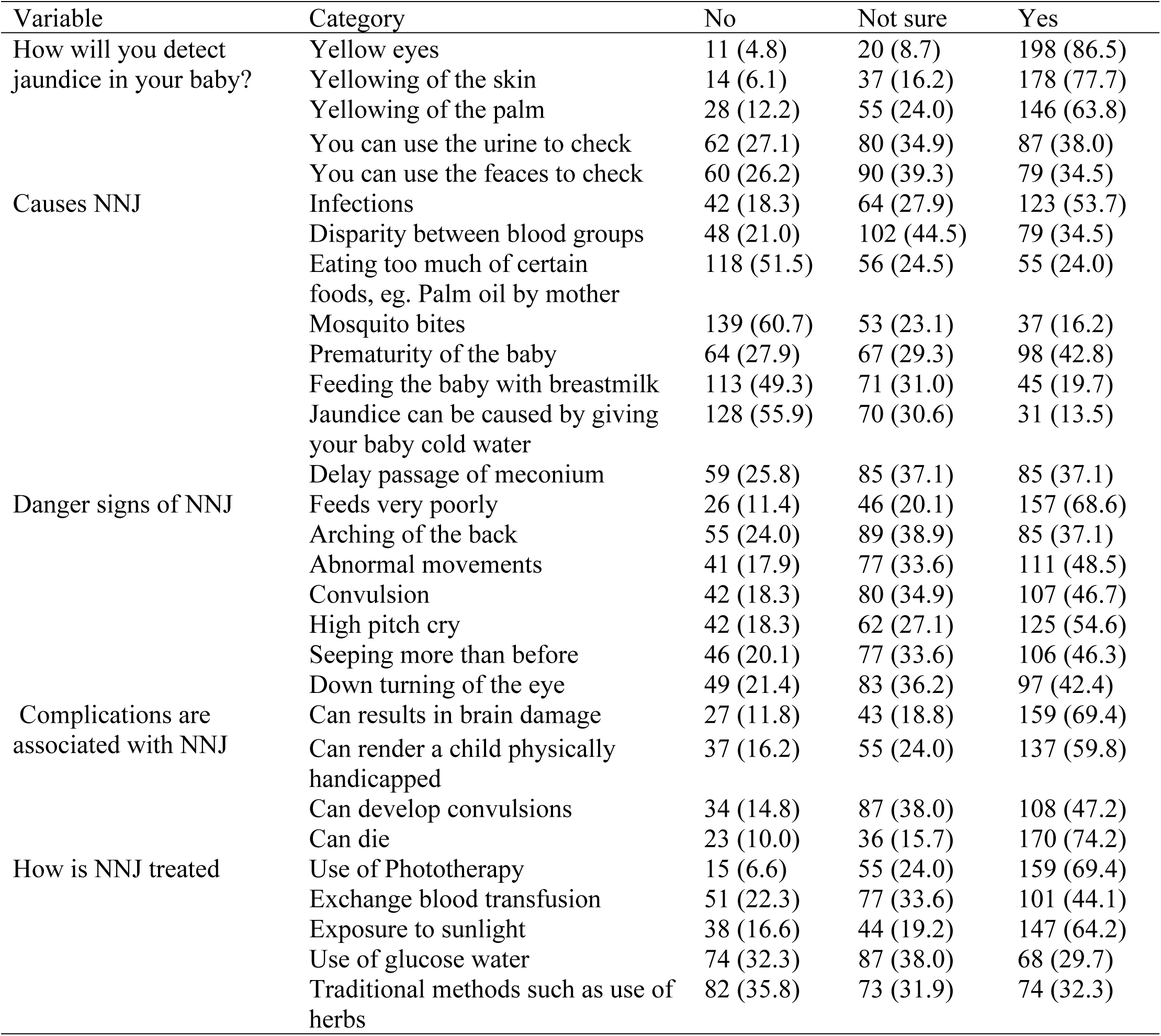
Mothers’ Knowledge Regarding NNJ.

Most mothers (65.1%) demonstrated a good attitude toward neonatal jaundice. Interestingly, nearly three-quarters (74.7%) agreed that attending antenatal care frequently equips them with knowledge to prevent and recognize the condition. While 78.2% acknowledged that jaundice can be treated at the hospital, a notable proportion (25.3%) still believed in herbal remedies. Furthermore, 56.8% endorsed exposing babies to early morning sunlight as a treatment, and 49.3% saw breastfeeding as beneficial. On the contrary, 66.8% dismissed the belief that evil spirits cause jaundice, and 59.8% disagreed with prayer or fasting as a preventive method (Table 4).

**Table 4:**
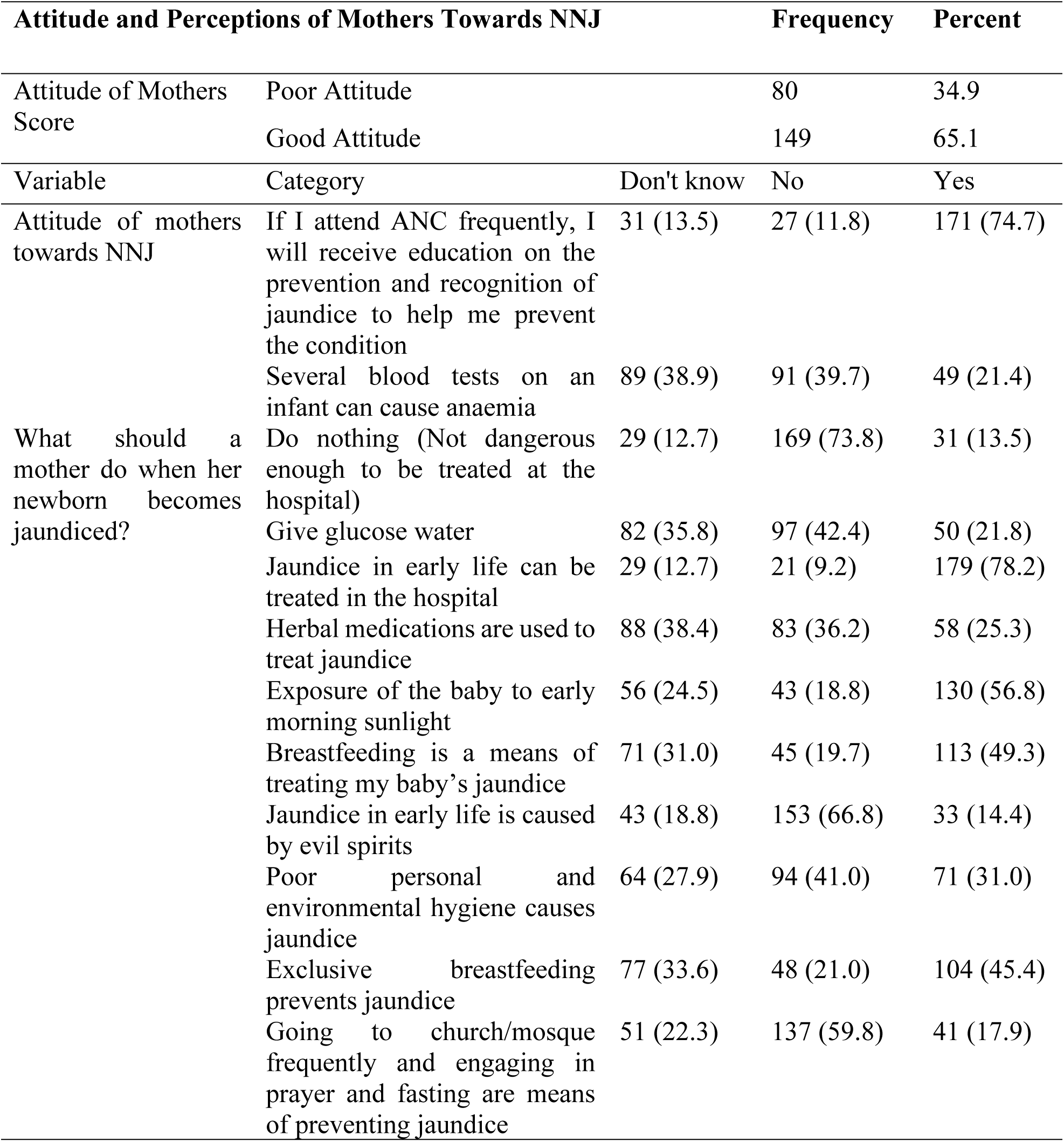
Attitude and Perceptions of Mothers Towards NNJ.

| Attitude and Perceptions of Mothers Towards NNJ |  |  | Frequency | Percent |
| --- | --- | --- | --- | --- |
| Attitude of Mothers Score | Poor Attitude |  | 80 | 34.9 |
|  | Good Attitude |  | 149 | 65.1 |
| Variable | Category | Don't know | No | Yes |
| Attitude of mothers towards NNJ | If I attend ANC frequently, I will receive education on the prevention and recognition of jaundice to help me prevent the condition | 31 (13.5) | 27 (11.8) | 171 (74.7) |
|  | Several blood tests on an infant can cause anaemia | 89 (38.9) | 91 (39.7) | 49 (21.4) |
| What should a mother do when her newborn becomes jaundiced? | Do nothing (Not dangerous enough to be treated at the hospital) | 29 (12.7) | 169 (73.8) | 31 (13.5) |
|  | Give glucose water | 82 (35.8) | 97 (42.4) | 50 (21.8) |
|  | Jaundice in early life can be treated in the hospital | 29 (12.7) | 21 (9.2) | 179 (78.2) |
|  | Herbal medications are used to treat jaundice | 88 (38.4) | 83 (36.2) | 58 (25.3) |
|  | Exposure of the baby to early morning sunlight | 56 (24.5) | 43 (18.8) | 130 (56.8) |
|  | Breastfeeding is a means of treating my baby's jaundice | 71 (31.0) | 45 (19.7) | 113 (49.3) |
|  | Jaundice in early life is caused by evil spirits | 43 (18.8) | 153 (66.8) | 33 (14.4) |
|  | Poor personal and environmental hygiene causes jaundice | 64 (27.9) | 94 (41.0) | 71 (31.0) |
|  | Exclusive breastfeeding prevents jaundice | 77 (33.6) | 48 (21.0) | 104 (45.4) |
|  | Going to church/mosque frequently and engaging in prayer and fasting are means of preventing jaundice | 51 (22.3) | 137 (59.8) | 41 (17.9) |

A significant majority (72.1%) of mothers demonstrated good traditional beliefs and practices toward neonatal jaundice. Most notably, over three-quarters rejected common traditional misconceptions such as believing jaundice is a curse (73.8%), or that yellow skin indicates beauty or good health (81.7% and 79.9%, respectively). Likewise, traditional practices such as putting babies in dark rooms (74.2%) or avoiding colostrum (76.0%) were rejected by many. However, a concerning 21.9% still believed that breast milk dropped into the baby’s eyes could cure jaundice (Table 5).

**Table 5:**
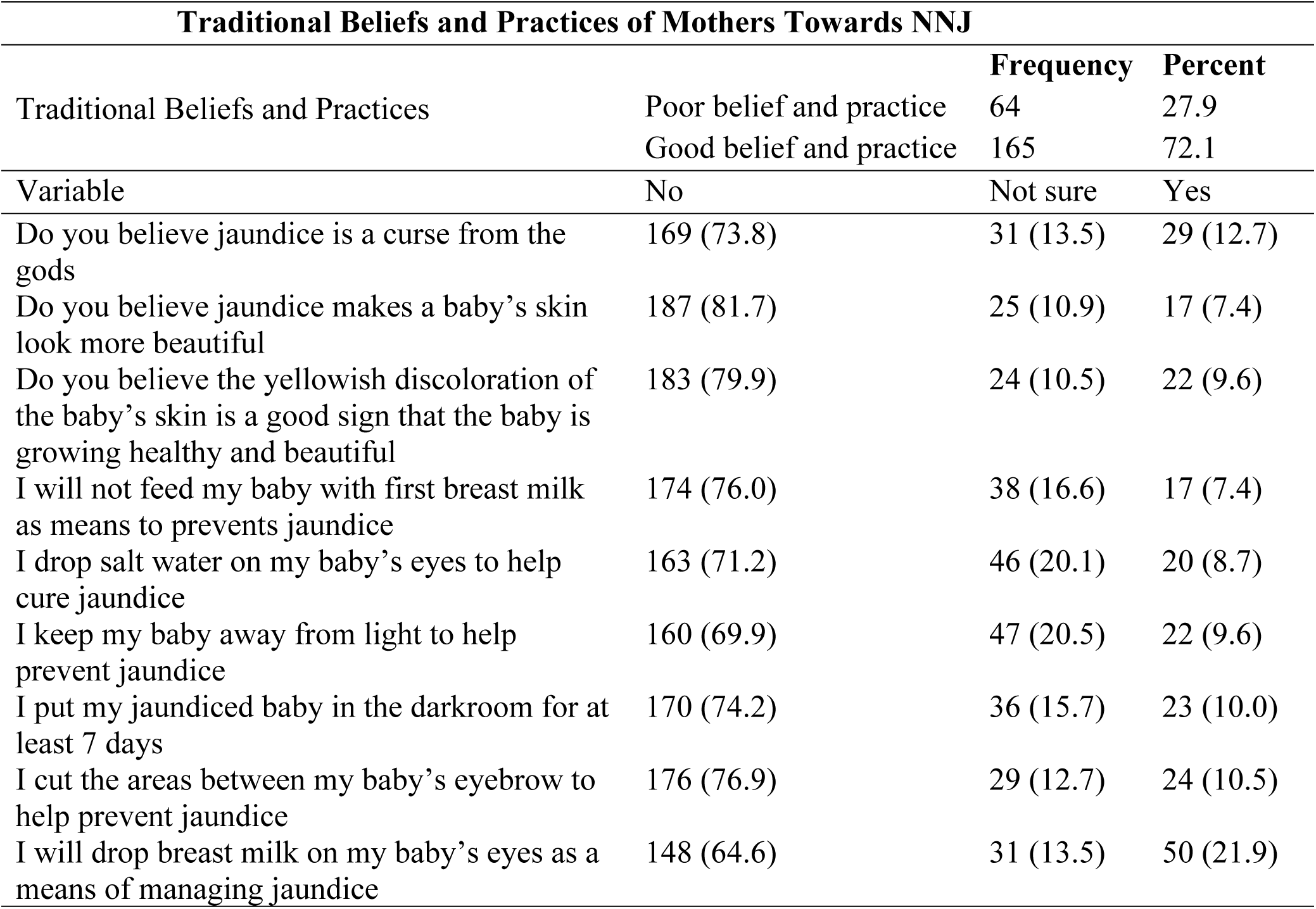
Traditional Beliefs and Practices of Mothers Towards NNJ.

The study revealed several predictors of mothers’ beliefs and practices towards NNJ (Table 6). Mothers with good knowledge of neonatal jaundice were over four times more likely to exhibit good traditional beliefs and practices compared to those with poor knowledge (AOR = 4.26; 95% CI: 1.68–10.79; p = 0.002). Additionally, those with a good attitude toward neonatal jaundice had significantly higher odds, with over nine times, of demonstrating appropriate beliefs and practices (AOR = 9.34; 95% CI: 3.72–23.42; p < 0.001). Ethnicity also showed a noteworthy association, as mothers from the Dagaba ethnic group were less likely to hold good beliefs and practices (AOR = 0.25; 95% CI: 0.07–0.95; p = 0.041). Unemployed mothers also had reduced odds of good beliefs and practices compared to their self-employed counterparts (AOR = 0.34; 95% CI: 0.13–0.90; p = 0.029).

**Table 6:** Determinants of Mothers’ Beliefs and Practices Towards NNJ.

| Variable | Poor belief and practice | Good belief and practice | X<sup>2</sup> (p-value) | COR (95% CI) p-value | AOR (95% CI) p-value |
| --- | --- | --- | --- | --- | --- |
| Age |  |  | 17.57 (<0.001) |  |  |
| 18-24 | 33 (44.0) | 42 (56.0) |  | Ref | Ref |
| 25-35 | 19 (16.4) | 97 (83.6) |  | 4.01 (2.05, 7.84) <0.001 | 2.10 (0.78, 5.63) 0.141 |
| 36-45 | 9 (31.0) | 20 (69.0) |  | 1.75 (0.70, 4.34) 0.230 | 0.49 (0.11, 2.22) 0.354 |
| 46+ | 3 (33.3) | 6 (66.7) |  | 1.57 (0.37, 6.76) 0.554 | 0.53 (0.03, 10.94) 0.681 |
| Marital status |  |  | 11.93 (0.008) |  |  |
| Single | 32 (40.5) | 47 (59.5) |  | Ref | Ref |
| Married | 24 (18.9) | 103 (81.1) |  | 2.92 (1.55, 5.50) <0.001 | 1.24 (0.36, 4.29) 0.731 |
| Divorced | 5 (33.3) | 10 (66.7) |  | 1.36 (0.43, 4.36) 0.603 | 1.37 (0.17, 10.97) 0.768 |
| Widowed | 3 (37.5) | 5 (62.5) |  | 1.13 (0.25, 5.09) 0.869 | 0.61 (0.04, 9.94) 0.726 |
| Sex |  |  | 3.22 (0.073) |  |  |
| Male | 0 (0.0) | 8 (100.0) |  |  |  |
| Female | 64 (29.0) | 157 (71.0) |  |  |  |
| Ethnicity |  |  | 14.24 (0.014) |  |  |
| Dagomba | 13 (17.1) | 63 (82.9) |  | Ref |  |
| Gonja | 9 (27.3) | 24 (72.7) |  | 0.55 (0.21, 1.45) 0.228 | 1.10 (0.30, 3.99) 0.886 |
| Akan | 10 (30.3) | 23 (69.7) |  | 0.47 (0.18, 1.23) 0.125 | 0.76 (0.196, 2.96) 0.694 |
| Dagaba | 11 (39.3) | 17 (60.7) |  | 0.32 (0.12, 0.84) 0.020 | 0.25 (0.07, 0.95) 0.041 |
| Frafra | 4 (17.4) | 19 (82.6) |  | 0.98 (0.29, 3.36) 0.975 | 0.62 (0.12, 3.33) 0.581 |
| Others | 17 (47.2) | 19 (52.8) |  | 0.23 (0.10, 0.56) <0.001 | 0.70 (0.16, 3.15) 0.644 |
| Religion |  |  | 5.86 (0.053) |  |  |
| Christians | 31 (28.7) | 77 (71.3) |  |  |  |
| Muslim | 27 (24.3) | 84 (75.7) |  |  |  |
| Traditionalist | 6 (60.0) | 4 (40.0) |  |  |  |
| Employment status |  |  | 17.62 (<0.001) |  |  |
| Self-employed/private | 20 (25.6) | 58 (74.4) |  | Ref | Ref |
| Unemployed | 38 (41.3) | 54 (58.7) |  | 0.49 (0.25, 0.94) 0.033 | 0.34 (0.13, 0.90) 0.029 |
| Public servant | 6 (10.2) | 53 (89.8) |  | 3.05 (1.14, 8.16) 0.027 | 2.00 (0.57, 7.05) 0.279 |
| Educational level |  |  | 7.71 (0.052) |  |  |
| None | 1 (11.1) | 8 (88.9) |  |  |  |
| Primary | 15 (46.9) | 17 (53.1) |  |  |  |
| Secondary | 20 (27.4) | 53 (72.6) |  |  |  |
| Tertiary | 28 (24.3) | 87 (75.7) |  |  |  |
| Parity |  |  | 13.11 (0.004) |  |  |
| 0 | 23 (46.0) | 27 (54.0) |  | Ref | Ref |
| 1 | 14 (26.4) | 39 (73.6) |  | 2.37 (1.04, 5.42) 0.040 | 0.73 (0.18, 3.08) 0.673 |
| 2 | 19 (26.8) | 52 (73.2) |  | 2.33 (1.09, 5.01) 0.030 | 0.36 (0.07, 1.84) 0.218 |
| >2 | 8 (14.5) | 47 (85.5) |  | 5.00 (1.97, 12.73) <0.001 | 0.81 (0.13, 5.11) 0.826 |
| Number of ANC visits |  |  | 1.35 (0.246) |  |  |
| <4 | 29 (32.2) | 61 (67.8) |  |  |  |
| 4+ | 35 (25.2) | 104 (74.8) |  |  |  |
| Knowledge | | | 39.45<br>($<0.001$) | | |
| Poor knowledge | 50 (48.5) | 53 (51.5) |  | Ref | Ref |
| Good knowledge | 14 (11.1) | 112 (88.9) | | 0.13 (0.07, 0.26) $<0.001$ | 4.26 (1.68, 10.79) 0.002 |
| Attitude | | | $<0.001$ | | |
| Poor attitude | 49 (61.3) | 31 (38.8) |  | Ref | Ref |
| Good attitude | 15 (10.1) | 134 (89.9) | | 14.12 (7.03, 28.38) $<0.001$ | 9.34 (3.72, 23.42) $<0.001$ |

## Discussion

This study found that just over half of the respondents (55.0%) had good knowledge of neonatal jaundice (NNJ), which, though modest, is slightly higher than figures reported in some earlier studies. For instance, Salia et al. in Ghana observed a lower knowledge rate of 45.5% [9], while Shrestha et al. in Nepal found that 59.9% of mothers had low knowledge regarding NNJ [17]. Likewise, Huq et al. [18] in Dhaka, Seneadza et al. [15] in Ghana, and Olatunde et al. [7] in Nigeria reported good knowledge among only 47.3%, 27.0%, and 30.5% of mothers, respectively. Compared to these, the current findings reflect a relative improvement, possibly due to increasing levels of formal education among mothers, enhanced antenatal education, and the vital role of health workers who were cited as the primary source of information by many respondents.

However, when compared to more encouraging findings such as Hameed et al. [1], who reported 71.5% good knowledge among participants, our results suggest that knowledge gaps still exist and merit further intervention. Mothers’ ability to recognize early signs like yellowing of the eyes and skin is promising, but misconceptions around traditional treatments, such as sunlight exposure and herbal remedies, persist. This may be attributed to intergenerational transfer of indigenous beliefs or lack of culturally sensitive education that directly addresses these misconceptions. Moreover, some mothers might confuse normal infant conditions with NNJ, resulting in delayed recognition and care-seeking behavior. Therefore, future health education strategies should not only deliver information but should also target and correct specific traditional beliefs and misunderstandings. On the attitudinal front, 65.1% of mothers demonstrated a good attitude towards NNJ, which aligns closely with findings from Al-Zamili and Saadoon, who reported 64% positive attitudes [5]. Similar patterns were noted by Huang et al. [19] in China and Salia et al. [9], both of whom found slightly lower figures (57.8% and 58.9% respectively). Positive attitudes are important as they often translate into health-seeking behaviours. Encouragingly, most mothers in this study rejected cultural myths such as NNJ being a curse, mirroring earlier findings by Salia et al. [9], where 84.7% rejected similar beliefs. This reflects an evolving understanding of the condition and suggests some success in public health campaigns and community-based sensitization efforts.

Traditional beliefs and practices in this study were largely appropriate, with 72.1% of mothers demonstrating good practices, higher than in Esan et al. [20], where only 60% of mothers would seek hospital care. The relatively higher rate in this study may be linked to increasing trust in health systems, accessibility of care, and prior experiences with childhood illnesses. However, pockets of harmful practices still exist. About 21.9% believed in applying breast milk to the eyes, and 56.8% supported sun exposure, reflecting trends seen in Egypt [21] and Bangladesh [18], where poor practices persisted despite education. These findings underscore the need for context-specific interventions that acknowledge cultural contexts while promoting safe newborn care practices.

Notably, this study identified several significant predictors of maternal beliefs and practices toward NNJ. Mothers with good knowledge of NNJ were more than four times as likely to exhibit appropriate beliefs and practices compared to those with poor knowledge. This supports the notion that awareness and comprehension of neonatal health conditions are foundational to positive caregiving practices. Furthermore, mothers who had a good attitude toward NNJ were over nine times more likely to exhibit appropriate beliefs and practices, reinforcing the critical role of emotional and perceptual factors in influencing health behaviours. These findings highlight the importance of integrated maternal education strategies that address both knowledge acquisition and attitudinal change.

The study was without limitation, as the cross-sectional design restricted the ability to establish causal relationships between knowledge, beliefs, and practices regarding neonatal jaundice. In addition, data were collected through self-reported responses, which may have introduced recall bias or social desirability bias, especially on sensitive or culturally influenced beliefs. Some mothers may have provided answers they perceived as acceptable to health professionals rather than their true practices. Lastly, language translation during interviews could have affected the interpretation of certain culturally nuanced terms, despite efforts to ensure clarity.

## Conclusion

The findings indicate that while a majority of mothers possessed good knowledge about neonatal jaundice, gaps remain in translating this knowledge into fully accurate beliefs and safe practices. Although many mothers could correctly identify symptoms, causes, and complications of neonatal jaundice, some continued to rely on outdated or potentially harmful practices such as exposing newborns to sunlight or applying breast milk to their eyes. This suggests that while health education has reached many mothers, it may not be sufficiently detailed or culturally sensitive to correct deep-rooted traditional misconceptions. Health promotion strategies should therefore be revised to include tailored, community-specific messaging delivered through trusted channels such as antenatal clinics and local media, emphasizing practical demonstrations and myth-busting education.

Moreover, while a substantial number of mothers rejected harmful traditional beliefs, a notable proportion still endorsed practices not supported by medical evidence. This highlights the persistent influence of cultural norms, suggesting that knowledge alone is not enough to effect behavioral change. To address this, interventions must incorporate community leaders, traditional birth attendants, and influencers in maternal health education efforts to bridge cultural and medical perspectives. Strengthening these collaborations will support mothers to make timely, informed decisions about neonatal health and reduce the risks associated with delayed recognition and treatment of jaundice.

## Data Availability

The data supporting the findings of this study are publicly available in the Figshare repository Link (https://figshare.com/s/77c9d7d893c4fdeac8e6) and can be accessed via the DOI: 10.6084/m9.figshare.29832659

## Acknowledgement

The authors sincerely acknowledge the healthcare workers at Tamale Teaching Hospital, Tamale West Hospital, and Tamale Central Health Center for their support during data collection. Special appreciation also goes to all the mothers who willingly participated and shared their experiences for the success of this study.

## References

[1] Hameed NN, Noor Abdul-Hussain FJ, Muna Ahmed MbcE, et al. Assessment of mother’s Knowledge, practices and believes toward home management of Neonatal jaundice in two pediatric teaching hospitals. J Fac Med Baghdad; 119. Epub ahead of print 2019. DOI: 10.32007/jfacmedbagdad.613,41710.

[2] Kassim NM, Obaid AF, Abdulrasol ZA. The Traditional Practices of Mothers in Caring of Neonates Affected by Hyperbilirubinemia. Iranian Rehabilitation Journal 2021; 19: 433– 440.

[3] Said N. Postnatal mother: Knowledge and attitude towards Neonatal Jaundice (NNJ). Elevate The International Journal of Nursing Education, Practice and Research 2018; 1: 40–45.

[4] Ng SY, Chong SY. What Do Mothers know about Neonatal Jaundice? Knowledge, Attitude and Practice of Mothers in Malaysia. Med J Malaysia; 69.

[5] Al-Zamili AH, Saadoon ZA. Knowledge, Attitude and Practice of Mothers to Neonatal Jaundice. Medico-legal Update; 20. Epub ahead of print March 2020. DOI: 10.37506/v20/i1/2020/mlu/194442.

[6] Boadi-Kusi SB, Holdbrook S, Kyei S, et al. Knowledge, Attitudes and Practices of Postnatal Mothers on Ophthalmia Neonatorum in the Central Region, Ghana. Health Serv Insights; 14. Epub ahead of print 2021. DOI: 10.1177/11786329211033248.

[7] Olatunde OE, Christianah OA, Olarinre BA, et al. Neonatal Jaundice: Perception of Pregnant Women Attending Antenatal Clinic at a Tertiary Hospital in Southwest, Nigeria. Glob Pediatr Health; 7. Epub ahead of print 2020. DOI: 10.1177/2333794X20982434.

[8] Ogunlesi TA, Abdul AR. Maternal knowledge and care-seeking behaviors for newborn jaundice in Sagamu, Southwest Nigeria. Niger J Clin Pract 2015; 18: 33–40.

[9] Salia SM, Afaya A, Wuni A, et al. Knowledge, attitudes and practices regarding neonatal jaundice among caregivers in a tertiary health facility in Ghana. PLoS One; 16. Epub ahead of print 1 June 2021. DOI: 10.1371/journal.pone.0251846.

[10] Abdul-Mumin A, Owusu EA, Wondoh PM, et al. Maternal Knowledge and Awareness of Neonatal Jaundice in Term Neonates admitted to the Neonatal Intensive Care Unit of the Tamale Teaching Hospital. Journal of Medical and Biomedical Sciences 2021; 8: 12–17.

[11] Goli H, Ansari M, Goli H, et al. Maternal experiences about neonatal jaundice: a qualitative study. Pediatric Anesthesia and Critical Care Journal 2020; 8: 58–64.

[12] Asiedu C, Edzeani EA. Assessing the Awareness and Perception of Neonatal Jaundice among Expectant Mothers in Ghana. J Pediatr Neonatal 2024; 6: 1–6.

[13] Ghafel HH, Al-Jubouri MB. Using unsafe traditional practices by Iraqi mothers to treat newborns’ problems. Heliyon; 10. Epub ahead of print 30 March 2024. DOI: 10.1016/j.heliyon.2024.e27842.

[14] Donkor DR, Ziblim SD, Dzantor EK, et al. Neonatal Jaundice Management: Knowledge, Attitude, and Practice Among Nurses and Midwives in the Northern Region, Ghana. SAGE Open Nurs; 9. Epub ahead of print 1 January 2023. DOI: 10.1177/23779608231187236.

[15] Seneadza NAH, Insaidoo G, Boye H, et al. Neonatal jaundice in Ghanaian children: Assessing maternal knowledge, attitude, and perceptions. PLoS One; 17. Epub ahead of print 1 March 2022. DOI: 10.1371/journal.pone.0264694.

[16] Demis A, Getie A, Wondmieneh A, et al. Knowledge on neonatal jaundice and its associated factors among mothers in northern Ethiopia: A facility-based cross-sectional study. BMJ Open; 11. Epub ahead of print 8 March 2021. DOI: 10.1136/bmjopen-2020-044390.

17. Shrestha S, Maharjan S, Petrini MA. Knowledge about Neonatal Jaundice among Nepalese Mothers. 2.

[18] Huq S, Hossain SM, Haque SMT, et al. Knowledge Regarding Neonatal Jaundice Management among Mothers: A Descriptive Study Done In a Tertiary Level Hospital of Dhaka City. Anwer Khan Modern Medical College Journal 2017; 8: 121–127.

[19] Huang Y, Chen L, Wang X, et al. Maternal knowledge, attitudes and practices related to neonatal jaundice and associated factors in Shenzhen, China: a facility-based cross-sectional study. BMJ Open; 12. Epub ahead of print 24 August 2022. DOI: 10.1136/bmjopen-2021-057981.

[20] Esan DT, Muhammad F, Ogunkorode A, et al. Traditional beliefs in the management and prevention of neonatal jaundice in Ado-Ekiti, Nigeria. Enfermería Clínica (English Edition) 2022; 32: S73–S76.

[21] Moawad EMI, Abdallah EAA, Ali YZA. Perceptions, practices, and traditional beliefs related to neonatal jaundice among Egyptian mothers A cross-sectional descriptive study. Medicine (United States); 95. Epub ahead of print 2016. DOI: 10.1097/MD.0000000000004804.

